# Reproducibility and transparency characteristics of oncology research evidence

**DOI:** 10.1101/19001917

**Authors:** Corbin G Walters, Zachery J Harter, Cole Wayant, Nam Vo, Michael Warren, Justin Chronister, Daniel Tritz, Matt Vassar

## Abstract

**Introduction:** As much as 50%-90% of research is estimated to be irreproducible, costing upwards of $28 billion in the United States alone. Reproducible research practices are essential to improving the reproducibility and transparency of biomedical research, such as including pre-registering studies, publishing a protocol, making research data and metadata publicly available, and publishing in open access journals. Here we report an investigation of key reproducible or transparent research practices in the published oncology literature.

**Methods:** We performed a cross-sectional analysis of a random sample of 300 oncology studies published from 2014-2018. We extracted key reproducibility and transparency characteristics in a duplicative fashion by blinded investigators using a pilot tested Google Form.

**Results:** Of the 300 studies randomly sampled, 296 studies were analyzed for study reproducibility characteristics. Of these 296 studies, 194 were contained empirical data that could be analyzed for reproducible and transparent research practices. Raw data was available for 9 studies (4.6%). Approximately 5 studies (2.6%) provided a protocol. Despite our sample including 15 clinical trials and 7 systematic reviews/meta-analyses, only 7 included a pre-registration statement. Less than 25% (65/194) of studies provided an author conflict of interest statement.

**Discussion:** We found that key reproducibility and transparency characteristics were absent from a random sample of published oncology studies. We recommend required pre-registration for all eligible trials and systematic reviews, published protocols for all manuscripts, and deposition of raw data and metadata in public repositories.

## Introduction

The ability to reproduce, or replicate, research results is a cornerstone of scientific advancement^1,2^. Absent efforts to advance the reproducibility of scientific research, advancements in patient care and outcomes may be delayed^3,4^, in part due to a failure in the translation of evidence to practice^5^. Evidence may fail translation to practice owing to bias^6,7^, lack of publication^4^, or poor reporting^8^. Thus, it may not be surprising that recent estimates of irreproducible research span a range of 50%-90% of all articles, costing upwards of $28 billion in the United States alone^9^. Moreover, it may not be surprising that large-scale efforts to replicate (ie, re-enact or reconduct previously published research studies) have failed^10^, in part due to an inability to navigate published methods. What is lost when scientific research fails to be reproducible carries significant weight; namely, the ability of science to be self-correcting^11^ and produce trustworthy results^12^.

It is commonly accepted that certain items are essential to improving the reproducibility of biomedical research. Examples of such items include pre-registering studies, publishing a protocol, making research data and metadata publicly available, and publishing in such a way to allow free access to the final manuscript. Pre-registering a study and publishing a protocol are important to prevent selective publication of studies with “positive” results^13^ and preventing the reordering of endpoints based on statistical significance^14,15^. Providing access to one’s raw research data, metadata, and analysis script allows independent researchers to computationally reproduce results, tailor results to specific patient populations, and determine the rigor of statistical analysis^16,17^. Publishing in open access journals or using preprint servers allows readers across economically diverse countries to access research articles that have implications for clinical practice^18^. Altogether, reproducible research practices aim to increase the efficiency, usefulness, and rigor of published research^5^.

Despite a high rate of author endorsement of reproducible practices^19,20^, some evidence suggests that authors infrequently implement them^21^. Absent such reproducible research practices, attempts to validate study findings may be thwarted. For example, Bayer and Amgen both attempted to replicate oncology research studies, with each failing to do so^22,23^. Bayer’s attempt to reproduce prior research studies is especially significant because they attempted to reproduce internal studies. Other non-pharmaceutical entities have attempted to replicate cancer research studies with similar results^24^. One may hypothesize that improved use and reporting of key reproducible or transparent research practices would improve future efforts to reproduce oncology research studies and build trust in existing evidence. Building on recent, similar analyses^25–27^, here we report an investigation of key reproducible or transparent research practices in the published oncology literature.

## Methods

We performed an observational study using a cross sectional design based on methods developed by Hardwicke et. al.^25^ with modifications. Our study employed best-practice design in accordance with published guidance, where relevant^28,29^. Study protocol, raw data, and other pertinent materials are available on the Open Science Framework (https://osf.io/x24n3/). This study did not meet U.S. regulation requirements to be classified as human research, therefore it is exempt from Institutional Review Board approval^30^.

### Journal Selection

We used the National Library of Medicine (NLM) catalog to search for all oncology journals using the subject terms tag Neoplasms[ST]. This search was performed on May, 29, 2019 which identified 344 journals. The inclusion criteria required that journals were both in “English” and “MEDLINE indexed”. We extracted electronic ISSN (or linking if electronic was unavailable)for each journal to use in a PubMed search on May 31, 2019. The total list of publications was then limited to those from January 1, 2014 to December 31, 2018.From search returns, we selected a random sample of 300 publications using Excel’s random number function (https://osf.io/wpev7/).

### Data Extraction

We used a pilot-tested Google Form based on the one provided by Hardwicke et. al. ^25^ with modifications (https://osf.io/3nfa5/). The first modifications were extracting the 5-year impact factor and the date of the most recent impact factor, neither of which were extracted by Hardwicke, et. al. Second, additional study designs were added to include cohort, case series, secondary analyses, chart reviews, and cross-sectional studies. Third, funding options were expanded that allowed for greater specification of university, hospital, public, private/industry, or non-profit sources.

The Google Form contained questions for investigators aimed at identifying whether a study demonstrated the information necessary to be reproducible (Table 1, Supplement 1). Variations in study design changed the data that was extracted from each study. For example, studies with no empirical data (e.g. editorials, commentaries [without reanalysis], simulations, news, reviews, and poems) were unable to examined for reproducibility characteristics. However, for all publications, the following data were extracted: title of study, 5 year impact factor, impact factor of the most recent year, country of corresponding author and publishing journal, type of study participants (eg, human or animal), study design, author conflicts of interest, funding source, and whether the article was open access (Table 2). Studies with empirical data were examined for the following characteristics in addition to those stated above: material and data availability, analysis scripts, linkable protocol, and trial pre-registration statements. Together, the 8 key reproducibility and transparency indicators analyzed were as follows: material availability, raw data availability, analysis scripts, linkable protocol, trial pre-registration statements, author conflict of interest statement, funding source, and open access. Open access was determined using www.openaccessbutton.org. In the event a study could not be found, investigators performed a Google search to see if the study was available elsewhere. Web of Science was used to evaluate whether each examined publication 1) had been replicated in other works and 2) was included in future systematic reviews or meta-analyses.

**Table 1:**
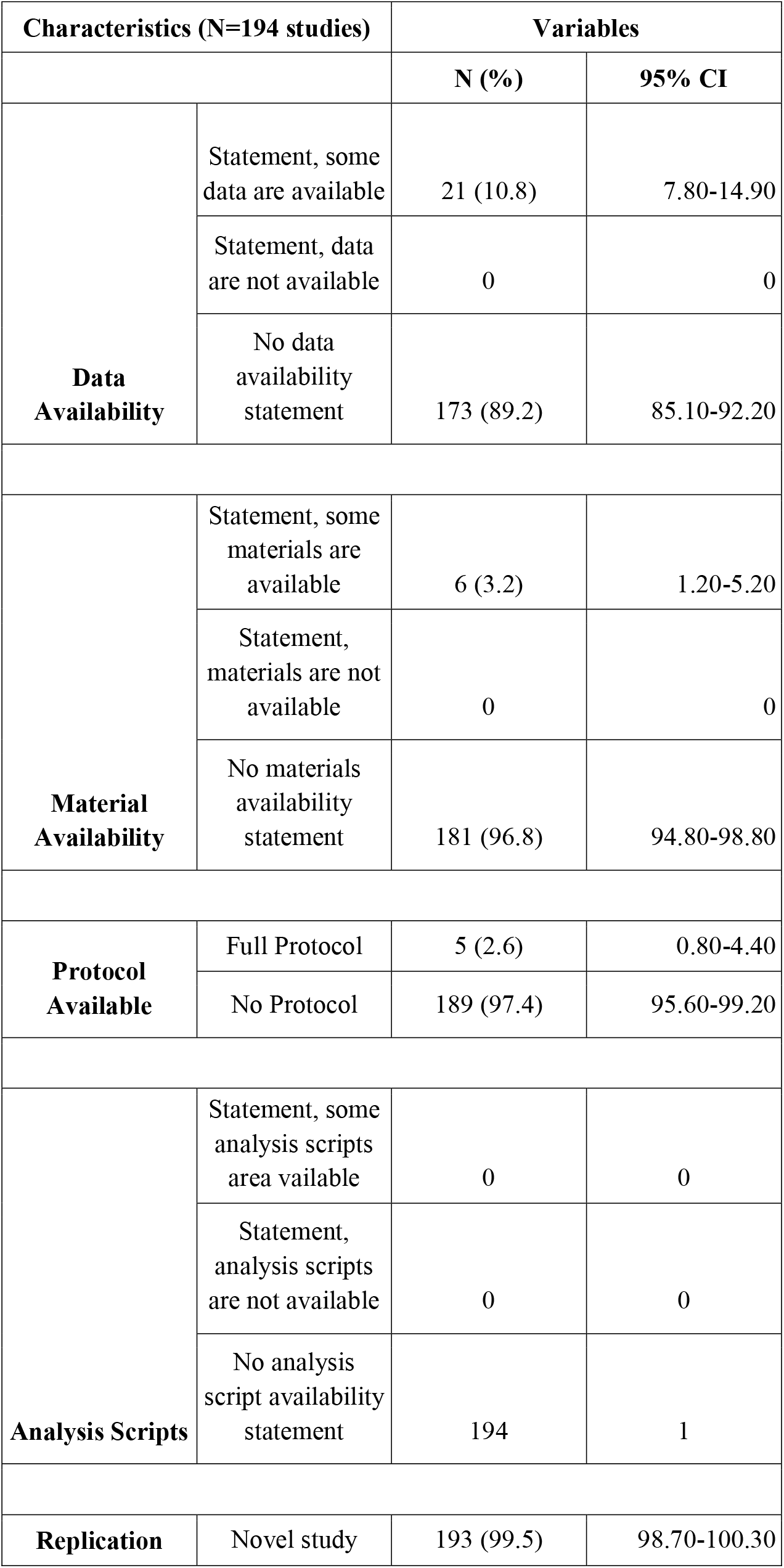
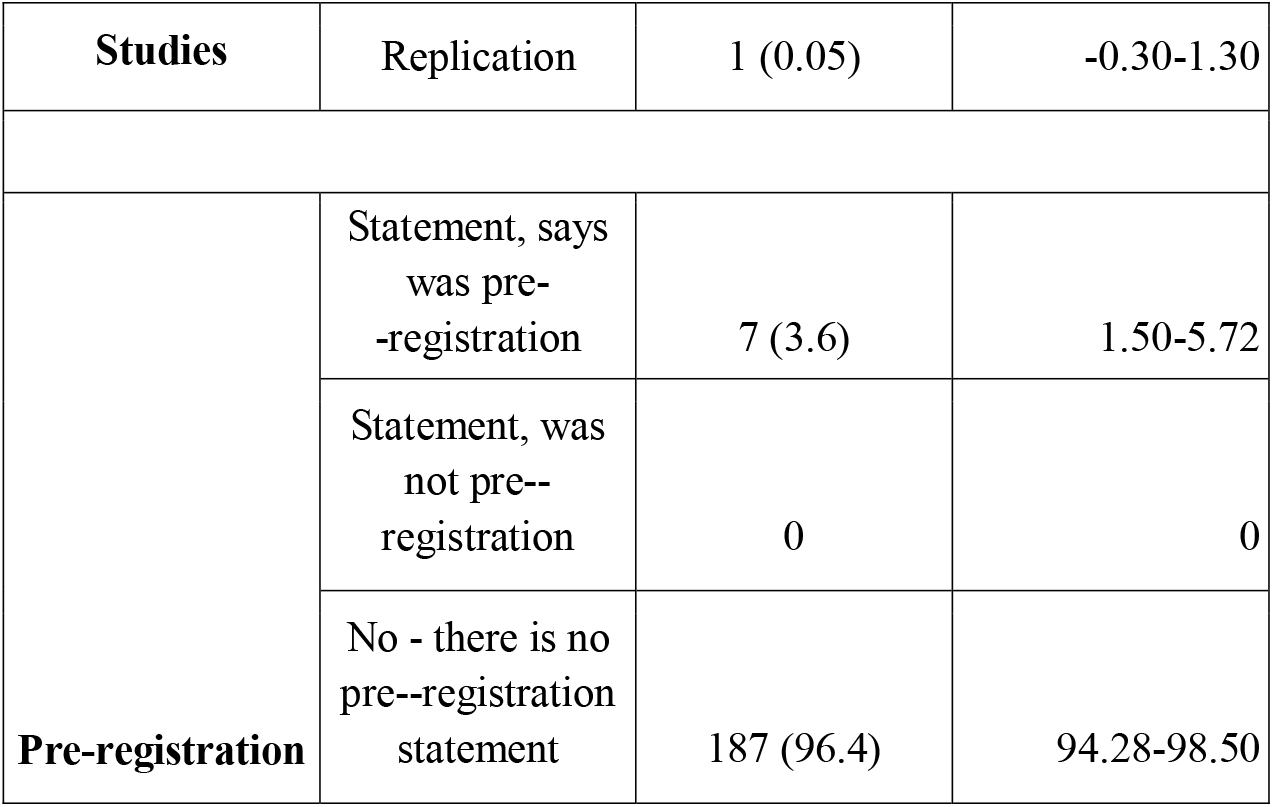
Reproducibility Characteristics of Oncology Studies.

**Table 2:** Characteristics of Oncology Studies.

| Table 2: Characteristics of Oncology Studies |  |  |  |
| --- | --- | --- | --- |
| Characteristics (N=296 studies) |  | Variables |  |
|  |  | N (%) | 95% CI |
| Test Subjects | Animals | 25 (8.5) | 5.30-11.60 |
|  | Humans | 154 (52.0) | 46.40-57.70 |
|  | Both | 0 | 0 |
|  | Neither | 117 (39.5) | 34.00-45.10 |
| Country of journal publication | US | 156 (52.7) | 47.05-58.35 |
|  | UK | 71 (24.0) | 19.15-28.82 |
|  | Greece | 18 (6.1) | 3.38-8.79 |
|  | Netherlands | 11 (3.7) | 1.58-5.86 |
|  | Ireland | 11 (3.7) | 1.58-5.86 |
|  | South Korea | 6 (2.0) | 0.43-3.62 |
|  | India | 4 (1.4) | 0.44-2.66 |
|  | Italy | 2 (0.7) | -0.25-1.60 |
|  | Japan | 2 (0.7) | -0.25-1.60 |
|  | Germany | 1 (0.3) | -0.32-0.99 |
|  | Unclear | 9 (3.0) | 1.97-4.98 |
|  | Other | 5 (1.7) | 0.23-3.15 |
| Country of corresponding author | US | 87 (29.4) | 24.24-34.55 |
|  | China | 52 (17.6) | 13.26-21.87 |
|  | Japan | 19 (6.4) | 3.65-9.19 |
|  | Germany | 16 (5.4) | 2.85-7.96 |
|  | South Korea | 13 (4.4) | 2.07-6.71 |
|  | UK | 12 (4.0) | 1.82-6.29 |
|  | Italy | 10 (3.4) | 1.33-5.42 |
|  | Canada | 7 (2.4) | 0.65-4.08 |
|  | France | 6 (2.0) | 0.43-3.62 |
|  | India | 6 (2.0) | 1.33-5.42 |
|  | Unclear | 8 (2.7) | 0.87-4.54 |
|  | Other | 60 (20.3) | 15.72-24.82 |
| **Funding** | University | 32 (10.8) | 7.30-14.30 |
|  | Hospital | 8 (2.7) | 0.90-4.50 |
|  | Public | 95 (32.1) | 26.80-37.40 |
|  | Private/Industry | 6 (2.0) | 0.40-3.60 |
|  | Non-profit | 7 (2.4) | 0.60-4.10 |
|  | No statement listed | 109 (36.8) | 31.40-42.30 |
|  | No funding recieved | 18 (6.1) | 3.40-8.80 |
|  | Mixed | 21 (7.1) | 4.19-10.0 |
| **Conflict of Interest statement** | Statement, one or more conflicts of interest | 57 (19.2) | 14.79-23.72 |
|  | Statement. no conflict of interest | 174 (58.8) | 53.21-64.35 |
|  | No conflict of interest statement | 65 (22.0) | 17.27-26.64 |
| **Open Access** | Yes - found via Open Access Button | 139 (47.0) | 40.69-51.97 |
|  | Yes - found article via other means | 26 (8.8) | 5.48-11.85 |
|  | No Could only access through paywall | 131 (44.2) | 38.05-49.28 |
| **5 Year Impact Factor** | Median | 3.445 | - |
|  | 1st quartile | 2.2705 | - |
|  | 3rd quartile | 5.95 | - |
|  | Interquartile range | 2.2705-5.95 | - |
| **Most Recent Impact Factor Year** | 2014 | 4 (1.4) | - |
|  | 2015 | 0 | - |
|  | 2016 | 4 (1.4) | - |
|  | 2017 | 271 (91.5) | - |
|  | 2018 | 1 (0.3) | - |
|  | Not Found | 16 (5.4) | - |
| **Most Recent Impact Factor** | Median | 3.346 | - |
|  | 1st quartile | 2.37375 | - |
|  | 3rd quartile | 6.471 | - |
|  | Interquartile range | 2.37375-6.471 | - |
| **Cited by Systematic Review N=296 (a)** | No Citations | 279 (94.3) | 91.60-96.90 |
|  | A Single Citation | 9 (3.0) | 1.10-5.00 |
|  | One to Five Citations | 8 (2.7) | 0.90-4.50 |
|  | Greater than Five Citations | 0 | 0 |
| **Cited by Meta-Analysis N=296 (b)** | No Citations | 274 (92.6) | 89.60-95.50 |
|  | A Single Citation | 13 (4.4) | 2.10-6.70 |
|  | One to Five Citations | 9 (3.0) | 1.10-5.00 |
|  | Greater than Five Citations | 0 | 0 |
| Abbreviations: CI, Confidence Interval. a - Two studies were explicitly excluded from the systematic reviews that cited the original article. b - Three studies were explicitly excluded from the meta-analysis that cited the original article |  |  |  |

Prior to data extraction, each investigator underwent a full day of training to increase the interrater reliability of the results between authors. This training consisted of an in-person session that reviewed study design, protocol, and Google Form. Investigators (C.G.W., N.V.) extracted data from 3 sample articles and differences were reconciled following extraction. A recording of this training session is available and listed online for reference (https://osf.io/tf7nw/). One investigator (C.G.W.) extracted data from all 300 publications. Z.J.H. extracted data for 200 publications and N.V. extracted data for 100 publications. C.G.W.’s data were compared to Z.J.H.’s and N.V.’s with discrepancies being resolved via group discussion. All authors were blinded to each other’s results. A final consensus meeting was held by all authors to resolve disagreements. If no agreement could be made, final judgment was made by an additional author (D.T.).

### Statistical Analysis

Descriptive statistics were calculated for each category using Microsoft Excel with 95% confidence intervals.

## Results

The NLM search identified 344 journals but only 204 fit our inclusion criteria. Our initial search string retrieved 199,420 oncology publications, from which, 300 were randomly sampled. Approximately 296 publications were analyzed for study reproducibility characteristics; 4 studies were not accessible, thus they were excluded from our analysis. Of these 296 publications, 215 contained empirical data and 81 did not. Publications without empirical data were unable to be analyzed for study reproducibility characteristics. Additionally, 21 publications with empirical data were case studies or case series. These case studies and series are unable to be replicated, thus are excluded from the analysis of study characteristics. In total, we were able to extract study reproducibility characteristics for 194 oncology publications (Figure 1).

### Study Characteristics

In our sample of oncology publications, the publishing journals had a median 5 year impact factor of 3.445 (IQR 2.27-5.95). The majority (156/296, 52.7%) of journals were located in the United States. Over half (165/296, 55.8%) of published studies were available for free via open access networks. The remaining 131 publications (44.2%) were located behind a paywall — making the studies inaccessible to the public — available only through paid reader access. Approximately 109 publications (36.8%) made no mention of funding source. Public funding (95/296, 32.1%), such as state or government institutions, comprised the next most prevalent source of study funding. Publication authors disclosed no conflict of interest more frequently than conflicts of interest (174/296, 58.8 vs. 57/296, 19.2%); however, 65 publications (22.0%) had no author conflict of interest statement. Human participants were the most common study population in sample (154/269, 52.0%). Citation rates of these 296 publications by systematic reviews and meta-analyses can be found in Table 2.

### Reproducibility Characteristics

Only 21 publications (21/194, 10.8%) made their raw data available. Approximately 9 of these publications with available raw data were downloadable by readers, while the rest was available upon request from the corresponding author of the study. A complete description of study materials required to reproduce the study — laboratory instruments, stimuli, computer software — was provided in 6/194 studies (3.2%). Of those publications with available materials, most were only accessible to readers upon request to the corresponding author, rather than being listed in a protocol or methods section. None of the included studies made their analysis scripts accessible, which details the steps the authors used to prepare the data for interpretation. Only 5 (5/194, 2.6%) publications provided a protocol detailing the *a priori* study design, methods, and analysis plan. Seven publications (7/194, 3.6%) were pre-registered in trial databases, such as ClinicalTrials.gov, prior to commencement of the study, despite there being 15 clinical trials and 7 systematic reviews/meta-analyses included in our analysis. One publication (1/194, 0.05%) claimed to be a replication study; all remaining studies (193/194, 99.5%) claimed to be novel or did not provide a clear statement about being a replication study. A subgroup analysis of the 8 key reproducibility and transparency indicators demonstrated that 29 publications had 0 indicators, 62 publications had 1 indicator, 209 publications had 2 to 5 indicators, and 0 publications had 6 or more.

## Discussion

Our cross-sectional investigation of a sample of the published oncology literature found that key reproducibility and transparency practices were lacking or entirely absent. Namely, we found that publications rarely pre-registered their methods, published their full protocol, or deposited raw data and analysis scripts into a publicly-accessible repository. Moreover, conflicts of interest were not discussed approximately 20% of the time and just over half of the included studies were not accessible due to journal paywalls. Given the challenges in understanding the molecular mechanisms that drive cancer, the continuum of research in the field of oncology is slow, laborious and inefficient^31^. To combat these inherent obstacles, transferring outcomes and information from preclinical to clinical research demands consistency and precision across the continuum. Otherwise, publications downstream in the cancer research continuum may be based on spurious results incapable of independent confirmation due to a lack of access to study data, protocols, or analysis scripts. Science advances more rapidly when people spend less time pursuing false leads^32^, thus, for patients with cancer and for whom rapid scientific advancement is most significant, it is paramount that scientists, researchers and physicians advocate for an efficient research system that is transparent, reproducible, and free from bias.

Pre-registration of research study methods is a mechanism to improve the reproducibility of published results and prevent bias — either from selective reporting of outcomes or selective publication of a study^33^. Previously, it has been shown that the selective reporting of study endpoints affects the research portfolio of drugs or diseases^15,34,35^. For example, Wayant et. al found that 109 RCTs of malignant hematology interventions selectively reported their trial endpoints 118 times, with a significant portion doing so in a manner that highlighted statistically significant findings^34^. Were trial registries not available, these trials may have never been found to exhibit selective outcome reporting. Now, through trial registries, hematologists and other interested researchers are able to independently assess the robustness of not only study rationale and results, but also study rigor and reporting. The present study indicates that pre-registration of study methods was rare, even among trials and systematic reviews that have available registries. The importance of preregistration across the continuum of cancer research cannot be understated. For example, preclinical animal models serve as the foundation for clinical trials, but have exhibited suboptimal methods^36^, which may explain why animal study results fail to successfully translate to clinical benefit. In fact, it was recently shown that many phase 3 trials in Oncology are conducted despite no significant phase 2 results^37^. One possible explanation for why phase 3 trials proceed despite nonsignificant phase 2 results is the strong bioplausibility demonstrated in preclinical studies. If it is true that preclinical studies exhibit poor research methods, it is not unlikely that they are affected by selective outcome reporting bias, just like clinical research studies. Thus, to strengthen oncology research evidence — from foundational, preclinical research to practice-changing trials — we recommend either the creation of relevant study registers or the adherence to existing registration policies. In so doing, one key aspect of research — the accurate reporting of planned study endpoints — could be monitored, detected, and mitigated.

Equally important to self-correcting, rigorous cancer research is the publication of protocols, raw data, and analysis scripts. Protocols include much more information than study outcomes — they may elaborate on statistical analysis plans or decisions fundamental to the critical appraisal of study results^38^. It is unlikely that anyone would be able to fully appraise a published study without access to a protocol, and far less likely that anyone would be capable of replicating the results independently. In fact, two recent efforts to reproduce preclinical studies revealed extant barriers to independent verification of published findings^20,39^, including the absence of protocols, data, and analysis scripts. Our present investigation found that only 5 (2.6%) studies published a protocol, 9 (4.6%) fully published their data, and none published their analysis scripts. In the context of the recent failures to reproduce cancer research studies, one may reasonably conclude that our study corroborates the belief that oncology research is not immune to the same shortcomings that contribute to an ever-expanding cohort of irreproducible research findings^40^. Oncology research, like all biomedical research, is at an inflection point, wherein it may progress toward more transparent, reproducible, efficient research findings. However, in order to do so, the availability of protocols, data, and analysis scripts should be considered fundamental.

In summary, we found that key reproducibility and transparency characteristics were absent from a random sample of published oncology studies. The implication of this finding is a research system that is incapable of rapid self-correction, or a research system that places a stronger emphasis on what is reported rather than what is correct. We recommend three key action items which we believe benefit oncology research and all its stakeholders. First, require pre-registration for eligible trials and systematic reviews, since these study designs have existing registries available, and support the development of registries for preclinical studies. Second, understand that published reports are snapshots of a research study, and require protocols be published. Last, encourage a scientific culture that relies on data that is true and robust, rather than author reports of their data, by requiring the deposition of raw data, meta data, and analysis scripts in public repositories.

This study has several strengths and limitations. First, for strengths, we sampled 300 published oncology articles indexed in PubMed. In doing so we captured a diverse array of research designs in an even more diverse range of journals. As such, all oncology researchers can read our paper and glean useful information and enact changes to improve the reproducibility of new evidence. With respect to our limitations, our study is too broad to make absolute judgments about specific study designs. All signals that suggest irreproducible research practices from our study fall in line with prior data in other areas of medicine^25–27^, but are nonetheless signals rather than answers. We suggest more narrow investigations of the reproducibility of specific study designs and suggest trials and animal studies be prioritized due to their potential influence (present or future) on patient care. Moreover, we do not suggest that irreproducible research findings are false; however, the trust in the results may be blunted. Further, replicating (ie, reconducting) a study is not necessary in all cases to assess the rigor of the results. If a protocol, statistical analysis plan, and raw data (including metadata) are available, one fundamental pillar of science would be reinforced: self-correction.

## Data Availability

Study protocol, raw data, and other pertinent materials are available on the Open Science Framework (https://osf.io/x24n3/).

## References

1. Munafò MR, Nosek BA, Bishop DVM, et al. A manifesto for reproducible science. Nature Human Behaviour. 2017;1(1):s41562-016-0021.

2. Peng R. The reproducibility crisis in science: A statistical counterattack. Significance. 2015;12(3):30–32.

3. Liberati A. An unfinished trip through uncertainties. BMJ. 2004;328(7438):531.

4. Scherer RW, Langenberg P, von Elm E. Full publication of results initially presented in abstracts. Cochrane Database Syst Rev. 2007;(2):MR000005.

5. Chalmers I, Glasziou P. Avoidable waste in the production and reporting of research evidence. Lancet. 2009;374(9683):86–89.

6. Hewitt C, Hahn S, Torgerson DJ, Watson J, Bland JM. Adequacy and reporting of allocation concealment: review of recent trials published in four general medical journals. BMJ. 2005;330(7499):1057–1058.

7. Whiting PF, Rutjes AWS, Westwood ME, Mallett S, QUADAS-2 Steering Group. A systematic review classifies sources of bias and variation in diagnostic test accuracy studies. J Clin Epidemiol. 2013;66(10):1093–1104.

8. Glasziou P, Meats E, Heneghan C, Shepperd S. What is missing from descriptions of treatment in trials and reviews? BMJ. 2008;336(7659):1472–1474.

9. Freedman LP, Cockburn IM, Simcoe TS. The Economics of Reproducibility in Preclinical Research. PLoS Biol. 2015;13(6):e1002165.

10. Kaiser J. Plan to replicate 50 high-impact cancer papers shrinks to just 18. Science. 2018;243.

11. Ioannidis JPA. Why Science Is Not Necessarily Self-Correcting. Perspect Psychol Sci. 2012;7(6):645–654.

12. Vazire S. Quality uncertainty erodes trust in science. Collabra: Psychology. 2017;3(1). https://collabra.org/articles/10.1525/collabra.74/print/.

13. Begg CB, Berlin JA. Publication bias and dissemination of clinical research. J Natl Cancer Inst. 1989;81(2):107–115.

14. You B, Gan HK, Pond G, Chen EX. Consistency in the analysis and reporting of primary end points in oncology randomized controlled trials from registration to publication: a systematic review. J Clin Oncol. 2012;30(2):210–216.

15. Mathieu S, Boutron I, Moher D, Altman DG, Ravaud P. Comparison of registered and published primary outcomes in randomized controlled trials. JAMA. 2009;302(9):977–984.

16. Voytek B. The Virtuous Cycle of a Data Ecosystem. PLoS Comput Biol. 2016;12(8):e1005037.

17. Steegen S, Tuerlinckx F, Gelman A, Vanpaemel W. Increasing Transparency Through a Multiverse Analysis. Perspect Psychol Sci. 2016;11(5):702–712.

18. Sarabipour S, Debat HJ, Emmott E, Burgess SJ, Schwessinger B, Hensel Z. On the value of preprints: An early career researcher perspective. PLoS Biol. 2019;17(2):e3000151.

19. Anderson MS, Ronning EA, Devries R, Martinson BC. Extending the Mertonian Norms: Scientists’ Subscription to Norms of Research. J Higher Educ. 2010;81(3):366–393.

20. Baker M. 1,500 scientists lift the lid on reproducibility. Nature. 2016;533(7604):452–454.

21. Harris JK, Johnson KJ, Carothers BJ, Combs TB, Luke DA, Wang X. Use of reproducible research practices in public health: A survey of public health analysts. PLoS One. 2018;13(9):e0202447.

22. Prinz F, Schlange T, Asadullah K. Believe it or not: how much can we rely on published data on potential drug targets? Nat Rev Drug Discov. 2011;10:712.

23. Begley CG, Ellis LM. Raise standards for preclinical cancer research. Nature. 2012;483(7391):531–533.

24. Errington TM, Iorns E, Gunn W, Tan FE, Lomax J, Nosek BA. Science forum: An open investigation of the reproducibility of cancer biology research. Elife. 2014;3:e04333.

25. Hardwicke TE, Wallach JD, Kidwell M, Ioannidis J. An empirical assessment of transparency and reproducibility-related research practices in the social sciences (2014-2017). April 2019. doi:10.31222/osf.io/6uhg5

26. Iqbal SA, Wallach JD, Khoury MJ, Schully SD, Ioannidis JPA. Reproducible Research Practices and Transparency across the Biomedical Literature. PLoS Biol. 2016;14(1):e1002333.

27. Wallach JD, Boyack KW, Ioannidis JPA. Reproducible research practices, transparency, and open access data in the biomedical literature, 2015–2017. PLoS Biol. 2018;16(11):e2006930.

28. Murad MH, Wang Z. Guidelines for reporting meta-epidemiological methodology research. Evid Based Med. 2017;22(4):139–142.

29. Moher D, Shamseer L, Clarke M, et al. Preferred reporting items for systematic review and meta-analysis protocols (PRISMA-P) 2015 statement. Syst Rev. 2015;4:1.

30. Office for Human Research Protections (OHRP). 45 CFR 46. HHS.gov. https://www.hhs.gov/ohrp/regulations-and-policy/regulations/45-cfr-46/index.html. Published February 16, 2016. Accessed July 10, 2018.

31. Celis JE, Pavalkis D. A mission-oriented approach to cancer in Europe: a joint mission/vision 2030. Mol Oncol. 2017;11(12):1661–1672.

32. Nuzzo R. How scientists fool themselves – and how they can stop. Nature. 2015;526(7572):182–185. doi:10.1038/526182a

33. Kaplan RM, Irvin VL. Likelihood of Null Effects of Large NHLBI Clinical Trials Has Increased over Time. PLoS One. 2015;10(8):e0132382.

34. Wayant C, Scheckel C, Hicks C, et al. Evidence of selective reporting bias in hematology journals: A systematic review. PLoS One. 2017;12(6):e0178379.

35. Chan A-W, Hróbjartsson A, Haahr MT, Gøtzsche PC, Altman DG. Empirical evidence for selective reporting of outcomes in randomized trials: comparison of protocols to published articles. JAMA. 2004;291(20):2457–2465.

36. Bara M, Joffe AR. The methodological quality of animal research in critical care: the public face of science. Ann Intensive Care. 2014;4:26.

37. Addeo A, Weiss GJ, Gyawali B. Association of Industry and Academic Sponsorship With Negative Phase 3 Oncology Trials and Reported Outcomes on Participant Survival: A Pooled Analysis. JAMA Netw Open. 2019;2(5):e193684.

38. Chan A-W, Hróbjartsson A. Promoting public access to clinical trial protocols: challenges and recommendations. Trials. 2018;19(1):116.

39. Collaboration O. Reproducibility Project: Cancer Biology. eLife. https://elifesciences.org/collections/9b1e83d1/reproducibility-project-cancer-biology. Accessed November 21, 2017.

40. Ioannidis JPA. Why most published research findings are false. PLoS Med. 2005;2(8):e124.

41. Swaen GG, Teggeler O, van Amelsvoort LG. False positive outcomes and design characteristics in occupational cancer epidemiology studies. Int J Epidemiol. 2001;30(5):948–954.

